# Anti-Correlation of KLRG1 and PD-1 Expression in Human Tumor CD8 T Cells

**DOI:** 10.1101/2023.03.01.23286593

**Authors:** Steven A Greenberg

**Author notes:** SG has completed the ICMJE uniform disclosure form at www.icmje.org/coi_disclosure.pdf and declares: SG is a founder and consultant to Abcuro, Inc. and inventor of intellectual property pertaining to KLRG1 targeting, owned and managed by the Brigham and Women’s Hospital. There are no other relationships or activities that could appear to have influenced the submitted work. Data used is publicly available as described in Methods. No funding was used for this study.

## Abstract

Recently, combination checkpoint therapy of cancer has been recognized as producing additive as opposed to synergistic benefit due to in part to positively correlated effects. The potential for uncorrelated or negatively correlated therapies to produce true synergistic benefits has been noted. Whereas the inhibitory receptors PD-1, CTLA-4, TIM-3, LAG-3, and TIGIT have been collectively characterized as exhaustion receptors, another inhibitory receptor KLRG1 was historically characterized as a senescent receptor and received relatively little attention as a potential checkpoint inhibitor target. The anti-tumor effects of KLRG1 blockade has relatively recently been demonstrated in preclinical in vivo studies. Here, expression of the inhibitory receptors PD-1, CTLA-4, TIM-3, LAG-3, TIGIT, and KLRG1 was studied in publicly available gene expression datasets. Bulk RNA microarray and RNAseq, and single cell RNAseq data from healthy blood and tumor tissue samples were analyzed for Pearson correlation. CD8 T cell differentiation of memory T cells from the TEM to TEMRA states is characterized by PD-1/KLRG1 anti-correlation, with decreased PD-1 expression but increased KLRG1 expression. Single cell RNAseq analysis of tumor infiltrating CD8 T cells shows positive correlation of CTLA-4, TIM-3, LAG-3, and TIGIT with PD-1 but negative correlation of KLRG1 with PD-1. The anti-correlation of PD-1 and KLRG1 expression in human tumor infiltrating CD8 T cells suggests the potential for combination therapy supra-additive benefits of anti-PD-1 and anti-KLRG1 therapies.

## Introduction

Targeting of the T cell inhibitory receptors PD-1, CTLA-4, TIM-3, LAG-3, and TIGIT for applications in oncology has gained widespread interest [1-3] and been tested in clinical studies. Therapeutic benefits in preclinical mouse studies with blocking antibodies was demonstrated for CTLA-4 by 1996 [4], PD-1 by 2005 [5, 6], LAG-3 by 2007 [7], TIM-3 by 2013 [8], and TIGIT by 2014 [9].

Other inhibitory receptors, such as KLRG1, have received little interest. A benefit for KLRG1 blockade was demonstrated in 2019 and again in 2021 [10, 11], with KLRG1 and PD-1 blockade producing supra-additive benefits in the B16F10 mouse melanoma model.

Historically, the pursuit of additional T cell inhibitory receptors has been driven by attention to co-regulated expression. Thus, studies in 2007 of the molecular signature of mouse CD8+ T cells during chronic viral infection identified coordinate regulation of all inhibitory receptors studied (including PD-1, CTLA-4, and LAG-3) except for two, KLRG1 and KLRA9 [12]. Subsequent nearest neighbor correlation studies of this data identified KLRG1 as uniquely anti-correlated with PD-1 in mouse CD8+ T cells during chronic viral infection [13]. The coordinated expression of PD-1 with TIGIT [9] has also been emphasized.

Ultimately, a view emerged that receptors with coordinated expression to each other constituted a class of exhaustion receptors that were promising therapeutic targets while receptors with low or negative correlation to PD-1 might not be attractive targets. This view was further cemented by 2011 and 2015 with nomenclature for cancer CD8 T cells characterizing CTLA-4, PD-1, LAG-3, and TIM-3 as marking “exhausted” cells, whereas KLRG1 marked senescent cells [14, 15]. Alternative views as early as 2012 have noted the arbitrary distinction between exhaustion and senescence applied to these receptors, that human CD8 T cell inhibitory receptor surface protein expression may depend on differentiation and that KLRG1 protein surface expression is much higher in CD8 effector memory and effector populations than PD-1, CTLA-4, TIM-3, and LAG-3 [16, 17]. Here, we point out the anti-correlation of KLRG1 expression with PD-1 across the human CD8 T cell population and note the potential for combination therapies of anti-correlated combinations to produce clinical benefits greater than the sum of the benefits of the individual therapies [18], an effect that has been unrealized in the field of combination inhibitory receptor blocking therapies in the treatment of cancer.

## Materials and Methods

Public domain RNAseq gene expression data of bulk immune cell populations in normal blood (GSE107011)[19] was analyzed for pairwise Pearson correlation of PD-1 (PDCD1) with other T cell inhibitory receptors across the differentiated T cell subsets CD4 TEMRA, CD8 TEM and TEMRA, and γδ T cells, and in lung cancer tumor infiltrating lymphocytes (TILS) from dataset GSE99531. Correlation analysis was performed of single cell RNAseq gene expression from cancer tissue datasets used in a previously published analysis [10] GSE72046 (melanoma), GSE98638 (HCC), GSE103322 (HNSCC), and GSE89567 (astrocytoma) and newly analyzed comprehensive datasets GSE102575 (melanoma) [20], GSE108989 (colorectal cancer) [21], and GSE99254 (non-small cell lung cancer) [22]. CD8 T cells were annotated as those T cells with non-zero expression of CD8. Published graphical data was abstracted using https://apps.automeris.io/wpd/. Processed data underlying Figures and Table 1 are included in the Supplemental File.

**Table 1.** Pearson correlation coefficients of CD8 T cell inhibitory receptor expression with PD-1 across various single cell RNAseq datasets.

| Dataset | Cancer | Publication | # of CD8 T Cells | Correlation Coefficient $r$ with PD-1 | | | | |
| --- | --- | --- | --- | --- | --- | --- | --- | --- |
|  |  |  |  | CTLA-4 | TIM-3 | LAG-3 | TIGIT | KLRG1 |
| GSE120575 | Melanoma | [20] | 7,043 | 0.27 | 0.35 | 0.21 | 0.32 | -0.07 |
| GSE108989 | Colorectal | [21] | 6,856 | 0.12 | 0.24 | 0.08 | 0.18 | -0.07 |
| GSE99254 | NSCLC | [22] | 6,380 | 0.08 | 0.20 | 0.09 | 0.14 | -0.09 |
| GSE72056 | Melanoma | [24] | 1,224 | 0.26 | 0.35 | 0.22 | 0.28 | -0.14 |
| GSE98638 | HCC | [25] | 687 | 0.29 | 0.30 | 0.14 | 0.23 | -0.06 |
| GSE103322 | HNSCC | [26] | 635 | 0.21 | 0.26 | 0.14 | 0.25 | -0.02 |
| GSE89567 | Astrocytoma | [27] | 120 | 0.40 | 0.08 | 0.25 | 0.36 | -0.10 |

## Results

### Distinct expression of KLRG1 vs PD-1 in healthy human blood and tumor infiltrating differentiated T cells

Across the bulk highly differentiated T cell population in normal blood, KLRG1 gene expression was anti-correlated with PD-1 (r=-0.377) (**Figure 1A**). Previous authors have noted that KLRG1 protein surface expression by flow cytometry increases substantially in TEM and TEMRA subsets (75-80% of CD8+ T cells), while PD-1 expression decreases from TEM (30%) to TEMRA (10%) subsets [17] (**Figure 1B**).

**Figure 1.**
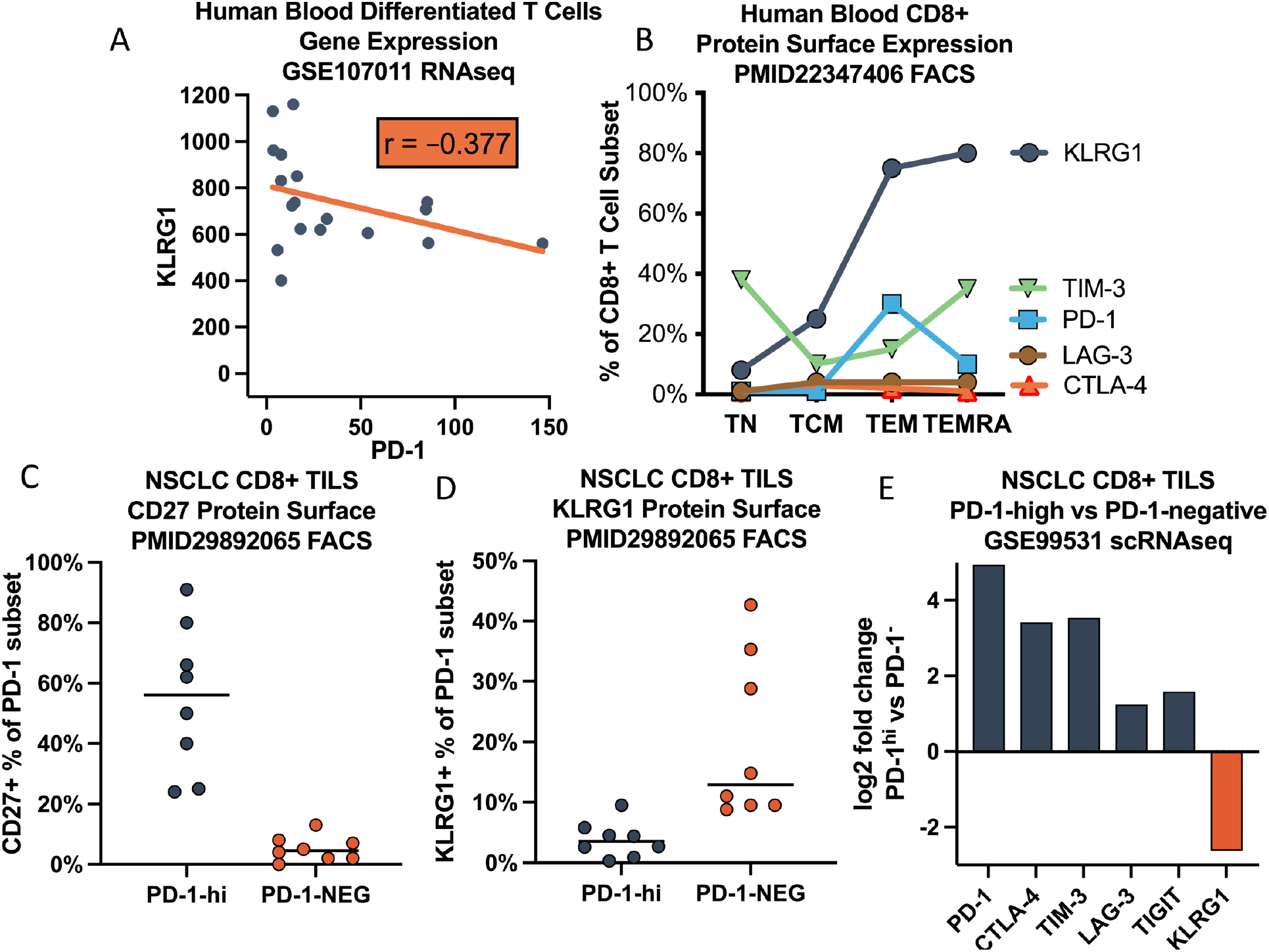
Distinct expression pattern of KLRG1 on highly differentiated human T cells. (A) Anticorrelated gene expression with PD-1 in RNAseq dataset GSE107011 for differentiated T cell subsets CD4 TEMRA, CD8 TEM, CD8 TEMRA, and γδ T cells. (B) Abstracted previously published data [17] for human blood CD8+ T cells protein surface expression showing increased KLRG1 on effector T (TEM) and effector (TEMRA) cells contrasting with decreasing PD-1 on TEMRA. (C-E) Characterization of PD-1+ CD8+ tumor infiltrating lymphocytes (TILS) in non-small cell lung cancer (NSCLC) datasets. (C) Abstracted data from previous publication [23] demonstrating that PD-1-high TILS are not highly differentiated TEMRA (CD27+) and (D) that KLRG1+ TILS are PD-1-. (E) Public domain single cell RNAseq (GSE99531) shows coregulated expression of CTLA-4, TIM-3, LAG-3, and TIGIT but anti-regulated KLRG1 expression in PD-1-high TILS.

A similar pattern has been reported [23] in non-small cell lung cancer (NSCLC) CD8+ tumor infiltrating lymphocytes (TILS), where PD-1-high cells are earlier stage CD27+ T cells, while KLRG1+ cells are later-stage CD27-T cells that are not PD-1-high (**Figure 1C**,**D**). More generally, NSCLC CD8+ PD-1-high vs PD-1-negative TILS have coregulated expression of CTLA-4, TIM-3, LAG-3, and TIGIT, but anti-regulated expression of KLRG1 (**Figure 1E**).

### Anti-correlation of KLRG1 and PD-1 expression across tumor infiltrating CD8+ T cells

To look more broadly at KLRG1 and PD-1 correlation, public domain single cell RNAseq gene expression datasets with available processed gene counts were obtained, and CD8 T cell data analyzed. Datasets of melanoma (N=7,043 CD8 T cells), colorectal cancer (N=6,856 CD8 T cells), non-small cell lung cancer (N=6,380 cells), and others showed positive correlation of CTLA-4, TIM-3, LAG-3, and TIGIT with PD-1 but anti-correlation of KLRG1 with PD-1 (**Figure 2; Table 1**).

**Figure 2.**
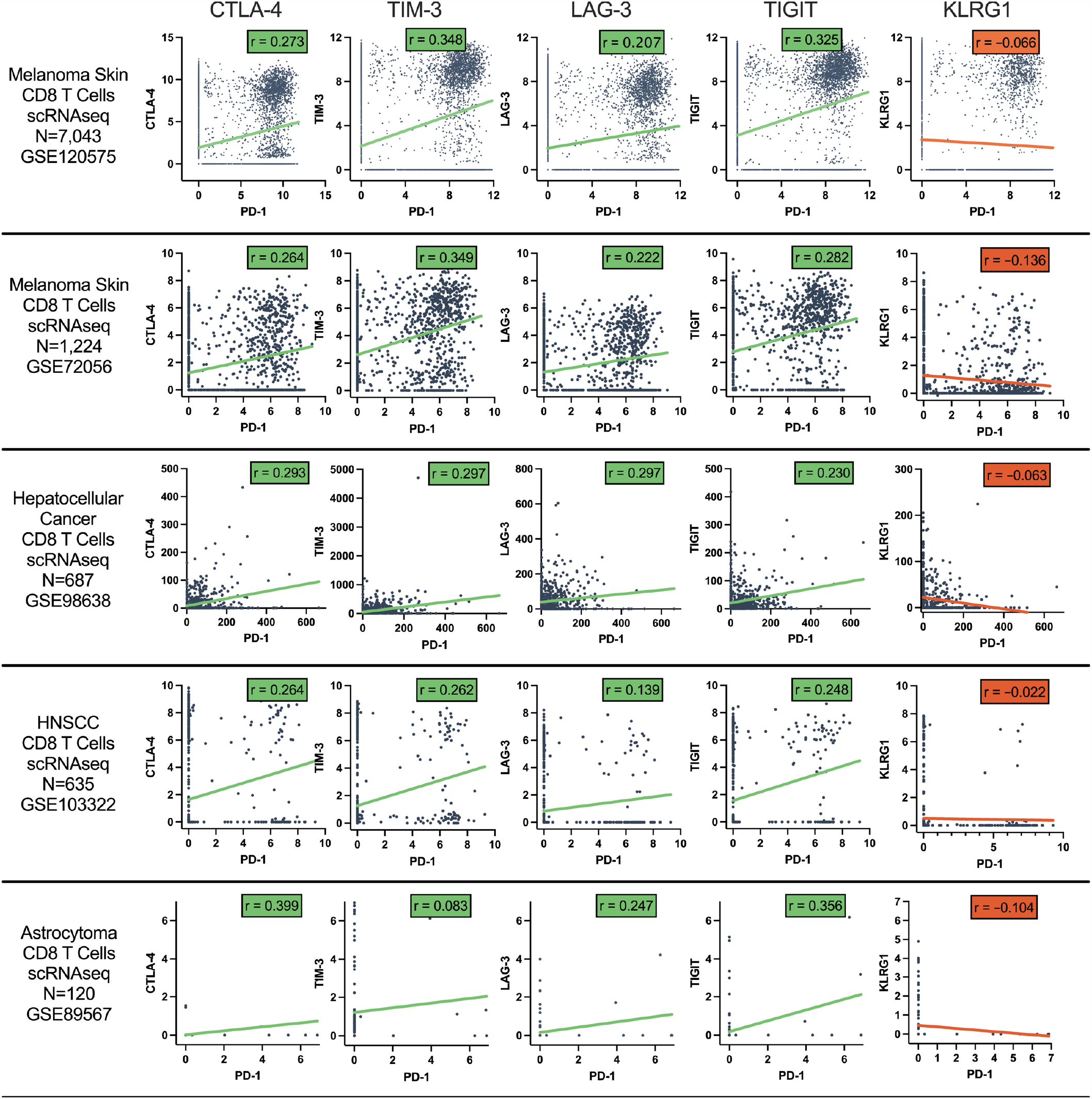
Pearson correlation of CD8 T cell co-inhibitory receptor gene expression with PD-1 across CD8 T cells from tumor samples. Data from single cell RNAseq datasets of melanoma, hepatocellular carcinoma (HCC), head and neck squamous cell cancer (HNSCC), and astrocytoma.

## Discussion

Much effort in the field of immuno-oncology has involved the study of combination therapies, including combinations involving blockade of more than one T cell inhibitory receptor. Combination therapies may produce clinical benefit through the additive independent action of each therapy or may produce supra-additive efficacy (in excess of that predicted by combined independent action) [28-30], which has been called clinical synergy [28]. Recent studies have demonstrated that the vast majority of combination studies with anti-PD-1 therapies have produced no more, and sometimes less, than the benefit expected from additive independent action [28, 31, 32].

Thus, supra-additive efficacy remains a desired goal. Although it can result from the well-established concept of pharmacological synergy, other mechanisms can also produce supra-additive efficacy [18]. For example collateral sensitivity (resistance to one drug confers susceptibility to another) can also produce clinical synergy, with antibiotic combinations used to overcome drug resistance a classic example. More generally, responses to combinations of monotherapies *a* and *b* are governed by

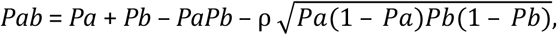

where *Pab* is the probability of response to the combination *ab* and ρ is the correlation coefficient of responses to *a* and *b* [18, 30]. Positively correlated therapies (ρ>0) can produce less than additive benefit and negatively correlated (anti-correlated) therapies (ρ<0) can produce supra-additive benefit.

Here, we point out that KLRG1 expression is concentrated on the most differentiated and potent CD8 T cells, unlike PD-1, and its expression is anti-correlated to PD-1 across CD8 T cells in a wide variety of cancers. Whereas much of the T cell inhibitory drug development efforts over the last decade have been focused on combinations of expression-correlated inhibitory receptor targets, the targeting of anti-correlated inhibitory receptors has greater potential to produce supra-additive benefit, and KLRG1 has this distinct property.

## Data Availability

All data used are publicly available in online databases and publications as described in Methods.

